# Cadmium Exposure and Hepato-renal Injury in Response to Cigarette Smoking in Apparently Healthy Active Smokers in Buea-Cameroon

**DOI:** 10.1101/2025.09.21.25336294

**Authors:** Nyambi Nkiackmenyi Ramin, Arnaud Fondjo Kouam, Elisabeth Menkem Zeuko’o, Armel Jackson Seukep, Armelle Gaelle Kwesseu Fepa, Fils Armand Ella, Pascal Dieudonné Chuisseu Djamen, Frédéric Nico Njayou, Paul Fewou Moundipa, Emmanuel Acha Asongalem

**Affiliations:** Department of Biomedical Sciences, Faculty of Health Sciences, University of Buea, PO Box 63, Buea, Cameroon; Department of Biochemistry, Faculty of Science, University of Yaoundé 1, P.O. Box 812, Yaounde 1, Cameroon; Higher Institute of Health Sciences, Université des Montagnes, P.O. Box 208, Bangangté, Cameroon

**Keywords:** Cigarette smoking, healthy active smokers, Cadmium exposure, Liver, Kidney, Injuries

## Abstract

Cigarette smoking in developing countries like Cameroon is increasing with unspecified exposure levels to cadmium, a known toxic chemical. This study conducted in Buea (Southwest Region, Cameroon) aimed to estimate the risk exposure to cadmium inhalation from smoking and its association with the abnormal level of liver and kidney function biomarkers in apparently healthy smokers. A survey recruited smokers to collect data on smoking habits. Cadmium contents in the major cigarette brands selected served to estimate the cadmium Daily Exposure (DE), Weekly Inhalational Exposure (WIE) and the percentage contribution to the Tolerable Weekly Intake (TWI) of cadmium. Serum alanine aminotransferase (ALT) activity, urea, creatinine contents, and urinary albuminuria and glucosuria were measured in blood and urine samples from active and nonsmokers. The 102 smokers enrolled were categorized respectively as light (37.3%; 38/102), moderate (38.2%; 39/102), and heavy (24.5%; 25/102) smokers, smoking an average number of 4.7 ± 0.5, 9.1 ± 1.5, and 19.1 ± 3.0 cigarettes/day, respectively. Cadmium levels in cigarette brands ranged from 0.72-1.08 µg Cd per gram of tobacco removed from the cigarette. Heavy smokers exceeded the permitted cadmium No Significant Risk Level (NSRL) by up to 28-fold, contributing 4-8% to the cadmium TWI. Serum ALT activity and creatinine content were significantly higher (*P*<0.05) in smokers groups compared to non-smokers. Also, moderate and heavy smokers were at a significantly higher risk (relative risk 6.90–13.32; *P*<0.05) of displaying abnormal elevated serum urea content, positive albuminuria and glucosuria. Our findings demonstrated that healthy active smokers are highly exposed to cadmium inhalation and are at higher risk of displaying abnormally increased levels of biochemical markers of liver and kidney functions, indicating risks of organ damage.

## Introduction

Environmental exposure to some chemicals poses a health concern and can cause damage to the body’s numerous systems and organs [1]. It is the instance of cadmium, a heavy metal widely present in food and plants, leading to universal exposure among individuals. Cadmium exposure is primarily through ingestion of water and food, inhaling industrial emissions, and smoking cigarettes for an extended period [2]. Indeed, cadmium is absorbed more effectively through inhalation than through ingestion; as a result, its content may increase four to five times in tobacco users compared to non-smokers [3].

Tobacco includes a variety of hazardous metals, cadmium being one of the most damaging [4,5]. This metal primarily penetrates tobacco plants (*Nicotina tabacum*) by absorption from the soil and is transported throughout the plant via zinc transporters [6,7]. Then cadmium accumulates naturally in tobacco leaves, which are the principal raw material utilized in cigarette manufacture. Accordingly, the amount of cadmium absorbed from smoking differs based on the brand of cigarette given that the cadmium content in one gram of dry tobacco leaf ranges from 1-2 µg, with a single cigarette containing about 0.5-1 µg of cadmium [8]. According to the World Health Organization (WHO), tobacco smoke is a serious public health hazard, accounting for over 8 million fatalities per year, with around 1.2 million non-smokers being exposed to secondhand smoke. More than 1.3 billion individuals worldwide consume tobacco products, with approximately 80% living in developing countries [9]. In Cameroon, tobacco smoking is estimated to be 5.3% in both urban and rural areas [10].

Cigarette smoking can harm almost every body’s organs and system. In addition to the lungs, cigarette smoking can damage the liver and kidneys, two important organs involved in the detoxification process of the organism [11]. Among the various toxic substances found in cigarettes, cadmium is well known for its toxicity to the kidneys and liver. Indeed, chronic exposure to cadmium leads to accumulation in these organs, causing cellular damage through oxidative stress and interference with normal cellular functions [12]. Toxins such as cadmium from smoking can cause oxidative stress, characterized by the overproduction of reactive oxygen species [13,14]. Sustained oxidative stress, due to persistent active smoking, can lead to liver injury and fibrosis, where tissue thickens and scars. Over time, fibrosis restricts blood flow and, in its severe form, becomes cirrhosis, a condition of permanent damage [15]. This ultimately decreases liver functions and promotes the development of hepatocellular carcinoma [16]. Cigarettes smoking can harm the kidney in multiple ways, including an increase in blood pressure, a reduction in renal blood flow, damage to glomeruli, and a gradual loss of kidney function [11,17]. In addition, smoking activity is a prominent risk factor for the progression of chronic kidney diseases, a state of irreversible kidney injury [18]. In the kidneys, cadmium primarily damages the proximal tubules, leading to the presence of protein or glucose in urine; and abnormal increased levels of blood urea and creatinine, two waste metabolic products excreted by the kidneys. The proteinuria or glucosuria as well as the abnormal serum level of creatinine and urea indicate compromised renal function, likely due to kidney damage [19]. In the liver, cadmium-induced oxidative stress can cause hepatocellular injury, reflected by elevated serum level of transaminases, including alanine aminotransferase [20]. Given the abovementioned harmful effects of cigarette smoking on the liver and kidney, early detection of biochemical indices reflecting signs of liver and kidney damage is necessary to enable a possible medical intervention while hepatic and renal function capacity have not yet been compromised.

Despite government advertising campaigns aim at discouraging people from smoking, and even the posters on cigarette packets clearly stating that “smoking kills”, the number of cigarette smokers in developing countries in general and Cameroon in particular, is growing alarmingly [21]. In addition, no comprehensive study has been undertaken to quantify the content of harmful chemicals such as cadmium found in cigarettes and to assess the effect of cigarette smoking on the hepatic and renal functions of apparently healthy active smokers. Considering these observations, this work aimed to evaluate the association of cigarette smoking with the abnormal level of liver and kidney function biomarkers of healthy active smokers, as well as estimate the level of cadmium risk exposure from smoking, based on the most popular brands of cigarettes smoked in the city of Buea, the Headquater of the Southwest, Cameroon.

## Materials and Methods

### Study area

This investigation was carried out in the city of Buea, the headquarter of South West Region of Cameroon. The city is located on the eastern slope of Mount Cameroon. It is characterized by an equatorial climate, with a temperature ranging between 18-35^°^C and two seasons: A long rainy season conveyed with high precipitation (up to 10,000 mm), which starts from the beginning of March till the end of October, and a short drying season, which spans from November to February. Buea is a university town, where the main professional activities are teaching and business (self-employment), with the vast majority of inhabitants being young adults (above 18 years). The area was therefore chosen to raise awareness among the young and old alike that smoking can potentially damage vital organs like the kidneys and the liver.

### Study design and target population

From May to October 2024, this cross-sectional study followed by laboratory investigations took-up consenting apparently active smokers (someone who smoked at least one cigarette per day during the last 12 months) aged 18 years and above. Febrile individuals as well as participants with history of liver or kidney diseases were excluded from the study.

### Sample size determination

Given that the primary aim of this study was to estimate the cadmium exposure level through cigarette smoking, the previous prevalence of cigarette smoking was considered in the estimation of sample size for of the study, which was calculated using Cochran’s Formula (1) [22].

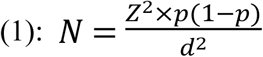

Where: *N* is the calculated sample size; *Z* = 1.96 is the typical value of the degree of confidence at 95%; *p* = 5.3% is the prevalence of cigarette smoking in Cameroon [23] (Njoumeni et al., 2020); *d* = 5% is the margin error permitted.

After calculation, *N* ≈ 78. For a better representation of the population, 20% of *N* was added. Therefore, the minimum number of apparently healthy active smokers to be enrolled in this study was 95 participants. In addition to that, 37 healthy individuals with no sign of any diseases, and without a history of smoking were also recruited and served as the control group during the analysis of biochemical markers of liver and kidney functions. This allows us to have an idea on how smoking can influence the variation of these biomarkers, as compared to apparently healthy non-smoker individuals.

### Sampling technique

Participants were recruited through a convenient non-probabilistic sampling technique from public spaces and smoking areas in Buea. After approaching a potential participant, the study objectives, procedures, and potential risks and benefits were clearly explained; and only participants who gave their consent by signing the informed consent were admitted to the study.

### Ethical statement

The study was conducted in accordance with guidelines from the declarations of Helsinki. An Ethical Clearance for this survey was obtained from the Institutional Review Board of the Faculty of Health Sciences, University of Buea (Reference number: 2024/2423-02/UB/SG/IRB/FHS). In addition, an Administrative Authorization was issued by the Regional Health Office of the South West Region, Ministry of Public Health, Cameroon. The study participant consisted exclusively of individuals who provided written informed consent.

## Data collection procedure

### Identification of the commercial cigarette brands consumed in Buea

A pre-tested structured questionnaire (Additional information S1_file) filled out by all participants was used to identify commercially available cigarette brands consumed in Buea. Socio-demographic characteristics (gender, age, marital status, education, occupation, and income level), as well as smoking habits (preferred cigarette brands, smoking history, number of cigarettes smoked per day, user of electronic cigarettes) of all participants were captured. Based on their daily number of cigarette smoked (excluding electronic cigarettes usage), the participants were categorized as light ([1–5] cigarettes/daily), moderate (]5-10] cigarettes/daily), and heavy smokers (]10-20] cigarettes/daily). At the end of the interview, 5 mL of blood was collected from each consenting patient by venipuncture in a dry tube and centrifuged (3000 × g, 5 min, 4°C). The serum obtained was used for the measurement of some biochemical markers of liver and kidney function. Likewise, approximately 20 mL of urine sample collected in a sterile urine cup was used for the qualitative detection of albumin and glucose.

### Determination of cadmium content of the identified brand of cigarettes

The identified cigarette brands based on the responses from the participants of the study were purchased in three different batches of production per brand with a difference of at least two months between production dates. Cadmium content in each sample was quantified as previously described [12]. In brief, the glassware to be used was soaked for 24 h in a 10% nitric acid solution and rinsed using demineralized water. From each of the packets purchased, six sticks of cigarette were selected and processed separately. After removing the filter, the remaining part of the cigarette was weighed and mineralized with 10 mL of the mineralization solution containing 75% nitric acid and 36% hydrochloric acid at the ratio of 1:3; v/v. The flask containing the mixture was heated at 90°C in a reflux device for about 3-4 h till the solution became colorless. The mineralized solution was allowed to cool down, filtered through Whatman filter paper N°1, and transferred into a 50 mL volumetric flask. The volume of the filtrate was completed to 50 mL using demineralized water and stored at 4°C until use. The concentration of cadmium was determined using an Atomic Absorption Spectrophotometer (Burk Scientific, In., Fort Point., USA.). Prior to analysis, calibration solutions with known concentrations of cadmium (0.125, 0.25, 0.5, 1, 2.5, 5, and 10 µg/L) prepared in demineralized water were used to calibrate the spectrophotometer and determine the content of cadmium. Then, 25 mL of mineralized samples were loaded into the Air-Acetylene flame of the spectrophotometer, with a lamp current of 4 mA, in which vaporization and atomization took place. The optical density was read at 288.8 nm.

## Risk exposure assessment

### Estimation of cadmium Daily Exposure (DE) and Cadmium Daily Inhalational Exposure (DIE) based on smoking habits

The estimation of DE (µg Cd/day) DIE (µg) and DIE (µg Cd/kg b.w. per day) through the smoking habits of healthy active smokers was determined using formula (2) and (3) respectively, based on the presumption that 10% of the total cadmium content in the cigarette is inhaled when smoking [24]. Only four of the most consumed brands of cigarettes based on the responses from participants were used for the risk assessment

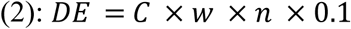

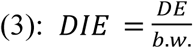

Where: *DE* is the cadmium Daily Exposure (µg Cd/day); *DIE* is the cadmium Daily Inhalational Exposure (µg Cd/kg b.w. per day); *C* is the concentration of cadmium of the cigarette brand (µg/g of cigarette); *w* is the weight of one cigarette (g); *n* is the number of cigarettes smoked per day; (for light, moderate, and heavy smokers, the number of cigarettes smoked per day were 5, 10, and 20 cigarettes, respectively). The factor 0.1 is the proportion of cadmium (10%) in cigarette assumed to be inhaled.

### Determination of fold difference between the estimated cadmium Daily Exposure and the permitted No Significant Risk Level (NSRL) of cadmium through inhalation

The fold difference comparison between the estimated cadmium daily exposure (DE) and the permitted No Significant Risk Level (NSRL) was determined as the ratio of DE value per smoker category and the NSRL value for cadmium (0.05 µg/day) as recommended by the California Environmental Protection Agency [25], using formula (4).

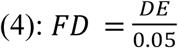

Where: *FD* is the fold difference between cadmium *DE* and *NSRL*; *DE* is the cadmium Daily Exposure (µg Cd/day); and 0.05 is the daily NSRL value for cadmium

### Estimation of Weekly Inhalational Exposure

Due to the bio-accumulative nature of cadmium in the body, a chronic exposure assessment is relevant to estimate the exposure to cadmium through cigarette smoking throughout the time. Accordingly, the Weekly Inhalational Exposure (WIE) to cadmium (µg Cd/kg. b.w. per week) per smoker category was determined using formula (5).

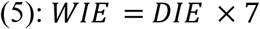

Where: WIE is the Weekly Inhalational Exposure (WIE) to cadmium (µg Cd/kg. b.w. per week); *DIE* is the cadmium Daily Inhalational Exposure (µg Cd/kg b.w. per day) and 7 is the number of day in one week.

### Determination of the percentage contribution of WIE to the Tolerable Weekly Cadmium Intake

The percentage contribution of cadmium through inhalation from cigarettes to the Tolerable Weekly Cadmium Intake (TWI) from all sources (cutaneous, ingestion, inhalation) was determined using formula (6).

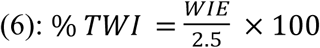

Where: %TWI is the percentage of the Tolerable Weekly Cadmium Intake; *WIE* is the cadmium Weekly Inhalational Exposure (µg Cd/kg. b.w. per week) and 2.5 µg Cd/kg b.w per week is the value of the total permitted weekly cadmium intake from all sources as recommended by the European Food Safety Authority [26].

### Measurement of biochemical markers of kidney and liver functions

The effects of cigarette smoking on the liver and kidney functions were assessed by measuring the serum activity of alanine aminotransferase (ALT), and serum contents of creatinine and urea using commercial assay kits (Catalog N° REF_ LP80507, Catalog N° REF_92314, and Catalog N° REF_80221 respectively for ALT, creatinine and urea) purchased from BIOLABO, Les Hautes Rives, Maizy, France. In addition, albuminuria and glucosuria were detected using Urine Test Strips/Uric 2V GP Glucose Protein Urine Analyzer (Catalog N° SG02102) purchased from Chongqing New World Trading Co., Ltd. China. All procedures were performed as recommended by the manufacturer’s instructions. For Urea and Creatinine, any value lower or higher than the upper limit of the normal range of values (ULN) was declared as normal or high, respectively. For ALT, any value lower or higher than 2 × ULN was considered normal or high, respectively. This classification was adapted from the RUCAM method (Roussel Uclaf Causality Assessment Method), which considers that variations in ALT values less than 2 × ULN could be due to individual variability and may not necessarily indicate liver damage [27,28].

### Data management and statistical analysis

After checking that all sections of the questionnaire had been completed, the data collected and the results of the laboratory analyses for each participant were saved in Excel 2016 (Microsoft Corporation, USA) (Additional information S2_file), and then exported to the statistical analysis software SPSS (Statistical Package for Social Sciences) version 25.0 (SPSS Inc., USA) or GraphPad Prism version 8.0.2. Descriptive statistics were performed using SPSS software. Qualitative variables were presented as frequency and percentage (%). Quantitative variables were first tested for normality using the Kolmogorov-Smirnov test. Variables that followed a normal distribution and variables that were abnormally distributed were expressed as mean ± standard deviation or median-interquartile range respectively. Quantitative data (median) between non-smokers and different smoker categories were compared by the non-parametric Student *t-test* followed by the Mann-Whitney *U* test. The statistical association between the different smoker categories and the levels (normal or abnormal) of liver and kidney function markers were determined using Fisher’s exact test, based on the contingency tables. Confounding variables were identified through bivariate and multivariate logistic regression analysis. The significance threshold was declared at *P*< 0.05.

## Results

### Socio-demographic characteristics of the enrolled apparently healthy active smokers

Table 1 presents the distribution of healthy active smokers participating in this study based on their socio-demographic traits. A total of 102 individuals were recruited with the majority being male, accounting for 93.1% (95/102). The mean age was 27.5 ± 5.1 years, with the age ranges]25-35] (72.5%; 74/102) and [18-25[(19.6%; 20/102) being the most common age groups. In terms of marital and education status, 88.2% of the participants (90/102) were unmarried, while 41.1% (42/102) had completed vocational training. 37.3% (38/102) were self-employed. 54.9% (56/102) of the participants had a lower-middle income, earning between 50,000 and 100,000 CFA francs monthly, which represents 100 to 200 US dollars per month.

**Table 1:** Socio-demographic characteristics of healthy active smokers enrolled in the study.

| **Socio-demographic characteristics** | **Category** | **Frequency (n)** | **Percentage (%)** |
| --- | --- | --- | --- |
| **Gender** | Female | 7 | 6.9 |
|  | Male | 95 | 93.1 |
|  | **Total** | **102** | **100.0** |
| **Age range** | [18-25] | 20 | 19.6 |
|  | ]25-35] | 74 | 72.5 |
|  | ]35-45] | 6 | 5.9 |
|  | ]45-55] | 2 | 2.0 |
|  | **Total** | **102** | **100.0** |
| **Marital status** | Married | 12 | 11.8 |
|  | Single | 90 | 88.2 |
|  | **Total** | **102** | **100.0** |
| **Education** | Primary | 2 | 2.0 |
|  | Secondary | 30 | 29.4 |
|  | Tertiary | 28 | 27.5 |
|  | Vocational | 42 | 41.1 |
|  | **Total** | **102** | **100.0** |
| **Occupation** | Civil servant | 2 | 2.0 |
|  | Manual labor | 32 | 31.3 |
|  | Self-employed | 38 | 37.3 |
|  | Unemployed | 30 | 29.4 |
|  | **Total** | **102** | **100.0** |
| **Income Level** | Low | 24 | 23.5 |
|  | Lower-middle | 56 | 54.9 |
|  | Middle | 22 | 21.6 |
|  | **Total** | **102** | **100.0** |

### Smoking habits of the studied population

Captured information from the participants on their smoking habits is summarized in Table 2.

**Table 2:** Distribution of healthy active smokers enrolled in the study according to their smoking habits.

| Smoking habits | Category | Frequency (n) | Percentage (%) |
| --- | --- | --- | --- |
| Number of years in smoking<br>(Year range) | [1-5] | 68 | 66.7 |
|  | ]5-10] | 24 | 23.5 |
|  | ]10-20] | 6 | 5.9 |
|  | >20 | 4 | 3.9 |
|  | **Total** | **102** | **100.0** |
| Number of cigarettes smoked per day<br>(Cigarette range) | [1-5] | 36 | 35.4 |
|  | ]5-10] | 40 | 39.2 |
|  | ]10-20] | 18 | 17.6 |
|  | >20 | 8 | 7.8 |
|  | **Total** | **102** | **100.0** |
| Smoker category | Light | 38 | 37.3 |
|  | Moderate | 39 | 38.2 |
|  | Heavy | 25 | 24.5 |
|  | **Total** | **102** | **100.0** |
| Use to electronic cigarette | No | 80 | 78.4 |
|  | Yes | 22 | 21.6 |
|  | **Total** | **102** | **100.0** |
| Alcohol consumption | No | 6 | 5.9 |
|  | Yes | 96 | 94.1 |
|  | **Total** | **102** | **100.0** |

Overall, most of the participants (66.7%; 68/102) were identified to have smoked for a duration between [1–5] years. The average daily number of cigarettes smoked was 10.6 ± 4.7 cigarettes/day and the most represented cigarette range group was]5-10] cigarettes/day (39.2%; 40/102). Concerning the smoker categories, 37.3% (38/102), 38.2% (39/102), and 24.5% (25/102) of the participants were classified respectively as: light smokers (smoking maximum 5 cigarettes/day), moderate smokers (smoking between 6-10 cigarette/day), and heavy smokers (smoking more than 11 cigarettes/days). In addition, nearly 1/4th of the responders declared themselves as users of electronic cigarettes (21.6%; 22/102) while the vast majority (94.1%; 96/102) consumed alcohol.

### Commercial brands of cigarettes identified from the survey

Table 3 presents the various brands of cigarettes consumed by healthy active smokers in Buea. A total of 11 imported brands of cigarettes from different origins were identified.

**Table 3:**
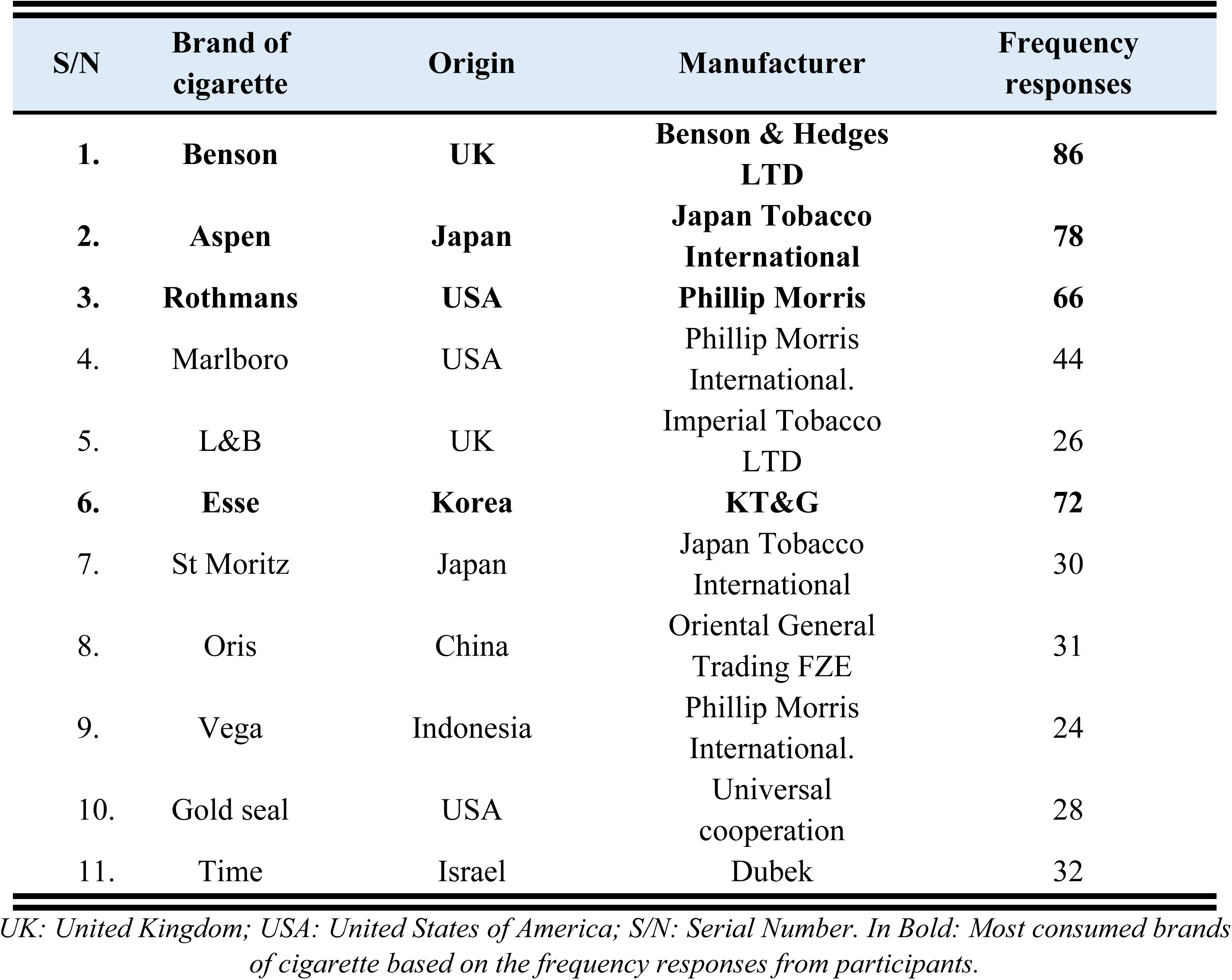
Commercial brand of cigarettes identified from the survey.

| S/N | Brand of cigarette | Origin | Manufacturer | Frequency responses |
| --- | --- | --- | --- | --- |
| 1. | **Benson** | **UK** | **Benson & Hedges LTD** | **86** |
| 2. | **Aspen** | **Japan** | **Japan Tobacco International** | **78** |
| 3. | **Rothmans** | **USA** | **Phillip Morris** | **66** |
| 4. | Marlboro | USA | Phillip Morris International. | 44 |
| 5. | L&B | UK | Imperial Tobacco LTD | 26 |
| 6. | **Esse** | **Korea** | **KT&G** | **72** |
| 7. | St Moritz | Japan | Japan Tobacco International | 30 |
| 8. | Oris | China | Oriental General Trading FZE | 31 |
| 9. | Vega | Indonesia | Phillip Morris International. | 24 |
| 10. | Gold seal | USA | Universal cooperation | 28 |
| 11. | Time | Israel | Dubek | 32 |

*UK: United Kingdom; USA: United States of America; S/N: Serial Number. In Bold: Most consumed brands* *of cigarette based on the frequency responses from participants.*

The most common brand of cigarettes smoked was Benson, imported from the United Kingdom, and consumed by 86/102 of the participants. The other popular brands included Aspen (from Japan), Esse (from Korea), and Rothmans (from the USA), smoked by 78/102, 72/102, and 66/102 of participants, respectively. In addition, an observation was that most participants typically smoked multiple brands, depending on the availability, or the cost. Other brands identified were Marlboro (USA), L&B (UK), St Moritz (Japan), Oris (China), Vega (Indonesia), Gold Seal (USA), and Time (Israel).

### Cadmium content in cigarette brands consumed in Buea

Fig 1 presents the concentration of cadmium in the various brands of cigarettes identified. The highest (1.08 ± 0.09 µg Cd/g of cigarette) and the lowest (0.72 ± 0.07 µg Cd/g of cigarette) concentrations were found in the brands L&B and Marlboro, respectively. Regarding the most popular brands (Benson, Aspen, Esse, and Rothmans), their cadmium contents ranged from 0.91 ± 0.07 to 0.96 ± 0.12 µg Cd/g of cigarette. These four brands were selected for the risk exposure assessment due to their popularity.

**Fig 1:**
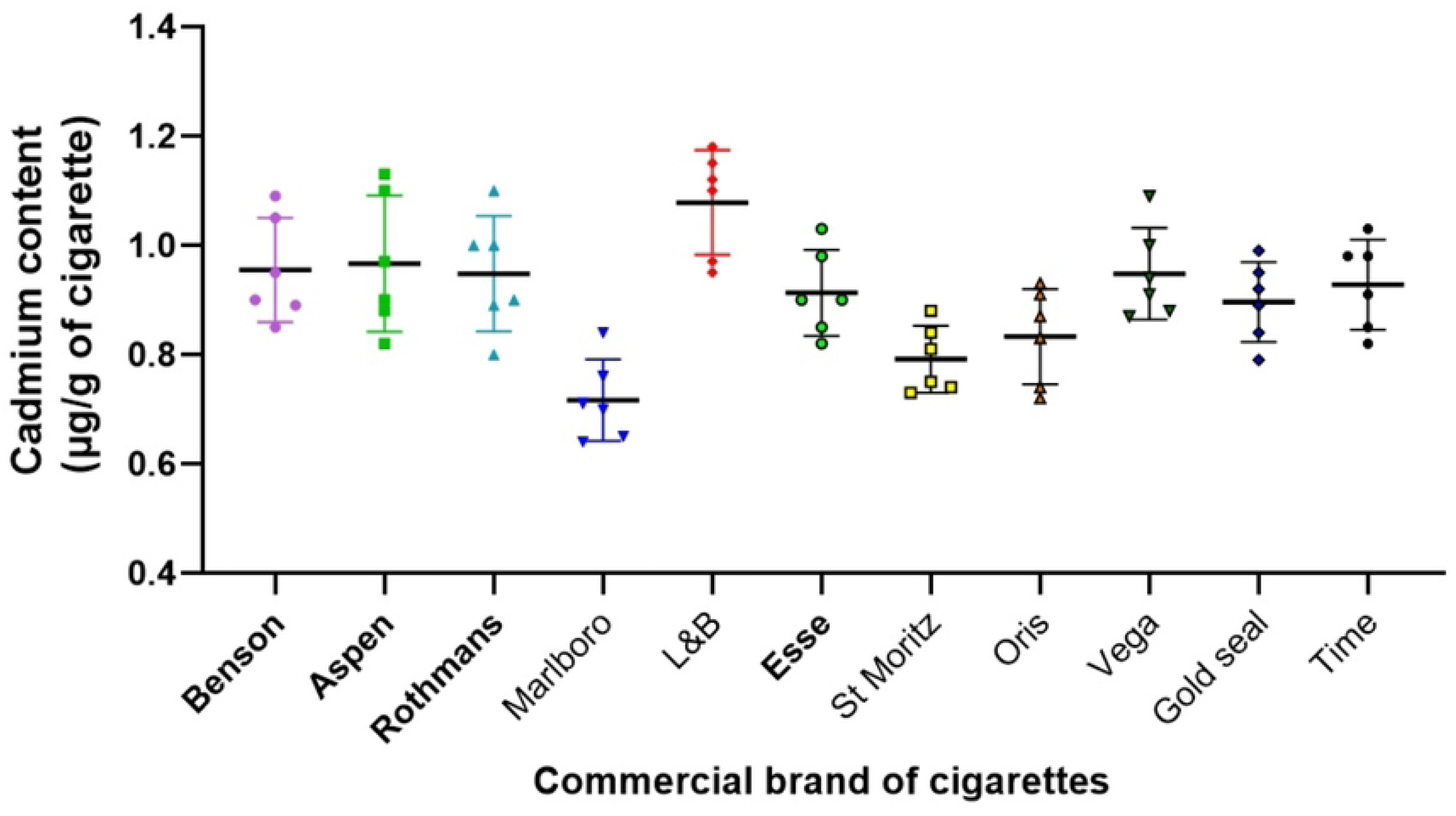
Concentration of cadmium of the identified commercial brand of cigarettes. Values are expressed as mean ± SD (n=6). The cigarette brand’s names written in bold are the most popular brands.

### Comparison of cadmium Daily Exposure from smoking with the permitted No Significant Risk Level – Inhalation of cadmium

The estimated cadmium Daily Exposure (DE, µg Cd/day) of the healthy active smokers enrolled in this study is presented in Table 4. It appears that the cadmium DE through smoking ranges from 0.18 to 1.44 µg Cd/day, depending on the smoking habits like the cigarette brand consumed, the smoker category, or the number of cigarettes consumed per day. These values were higher than the permitted NSRL **–** Inhalation value of cadmium (0.05 µg Cd/day). Indeed, the fold difference as compared to the permitted NSRL **–** Inhalation of cadmium for the estimated lowest value (brand: Esse; light smoker: 5 cigarettes/day) and the estimated highest value (brand: Benson; heavy smoker: 20 cigarettes/day) of the cadmium DE were 3 and 28, respectively.

**Table 4:** Mean value of DE estimates and fold difference to the permitted NSRL of Cadmium based on the smoker category and the most common brand of cigarette.

| Most common brand of cigarette | Smoker Category | Number of Cigarette/Day | Mean DE ($\mu\text{g Cd/day}$) | FD to the permitted NSRL |
| --- | --- | --- | --- | --- |
| **Benson** | Light | 5 | 0.36 | 7 |
|  | Moderate | 10 | 0.72 | 14 |
|  | Heavy | 20 | 1.44 | 28 |
| **Aspen** | Light | 5 | 0.225 | 4.5 |
|  | Moderate | 10 | 0.45 | 9 |
|  | Heavy | 20 | 0.9 | 18 |
| **Rothmans** | Light | 5 | 0.315 | 6 |
|  | Moderate | 10 | 0.63 | 12 |
|  | Heavy | 20 | 1.26 | 25 |
| **Esse** | Light | 5 | 0.18 | 3 |
|  | Moderate | 10 | 0.36 | 7 |
|  | Heavy | 20 | 0.72 | 14 |

*DE: Cadmium Daily Exposure ( $\mu\text{g Cd/day}$ ); FD: Fold Difference; NSRL: No Significant Risk Level* *– Inhalation of Cadmium; Permitted NSRL value of Cadmium: $0.05 \mu\text{g Cd/day}$ .*

### Contribution of the Cadmium Weekly Inhalational Exposure from smoking to the permitted Tolerable Weekly Intake of Cadmium

The estimated cadmium Weekly Inhalational Exposure (WIE, µg Cd/kg b.w. per week concerning various smoking habits and their percentage contribution to the permitted Tolerable Weekly Intake (%TWI) of cadmium are summarized in Table 5. For light and heavy smokers with an average body weight of 70kg, the cadmium WIE through smoking accounted for 1-2% and 4-8% of the TWI of cadmium, respectively.

**Table 5:** Mean values of DIE, WIE, and percentage contribution to the TWI of cadmium through smoking, based on the smoker category and the most common brand of cigarettes.

| Most common brand of cigarette | Smoker Category | Number of Cigarette/Day | Mean DIE ($\mu\text{g Cd/kg b.w./day}$) | Mean WIE ($\mu\text{g Cd/kg b.w./week}$) | %TWI |
| --- | --- | --- | --- | --- | --- |
| **Benson** | Light | 5 | 0.0072 | 0.0504 | 2.016 |
|  | Moderate | 10 | 0.0144 | 0.1008 | 4.032 |
|  | Heavy | 20 | 0.0288 | 0.2016 | 8.064 |
| **Aspen** | Light | 5 | 0.0045 | 0.0315 | 1.26 |
|  | Moderate | 10 | 0.009 | 0.063 | 2.52 |
|  | Heavy | 20 | 0.018 | 0.126 | 5.04 |
| **Rothmans** | Light | 5 | 0.0063 | 0.0441 | 1.764 |
|  | Moderate | 10 | 0.0126 | 0.0882 | 3.528 |
|  | Heavy | 20 | 0.0252 | 0.1764 | 7.056 |
| **Esse** | Light | 5 | 0.0036 | 0.0252 | 1.008 |
|  | Moderate | 10 | 0.0072 | 0.0504 | 2.016 |
|  | Heavy | 20 | 0.0144 | 0.1008 | 4.032 |

*DIE: Cadmium Daily Inhalational Exposure ( $\mu\text{g Cd/kg b.w./day}$ ); WIE: Cadmium Weekly* *Inhalational Exposure ( $\mu\text{g Cd/kg b.w./weeky}$ ); TWI: Tolerable Weekly Intake of Cadmium from all* *sources; Threshold TWI value of Cadmium: 2.5 $\mu\text{g Cd/kg b.w. per week}$ .*

### Influence of smoker categories on the variation of serum biochemical makers of liver and kidney injuries

Fig 2 illustrates the variation of serum ALT activity, serum urea, and creatinine contents according to the different smoker categories. The ALT activity in all smoker categories (light, moderate, or heavy) was significantly (*P*<0.05) abnormally elevated as compared to that of non-smokers (Fig 2A). Neither significant difference (*P*˃0.05) in urea content for light smokers (Fig 2B) nor in creatinine content (Fig 2C) for light and moderate smokers were found when compared to the non-smokers. Nevertheless, the serum content of urea for moderate and heavy smokers (Fig 2B) and the creatinine content for heavy smokers (Fig 2C) were significantly (*P*<0.05) elevated, as compared to the non-smokers, respectively.

**Fig 2:**
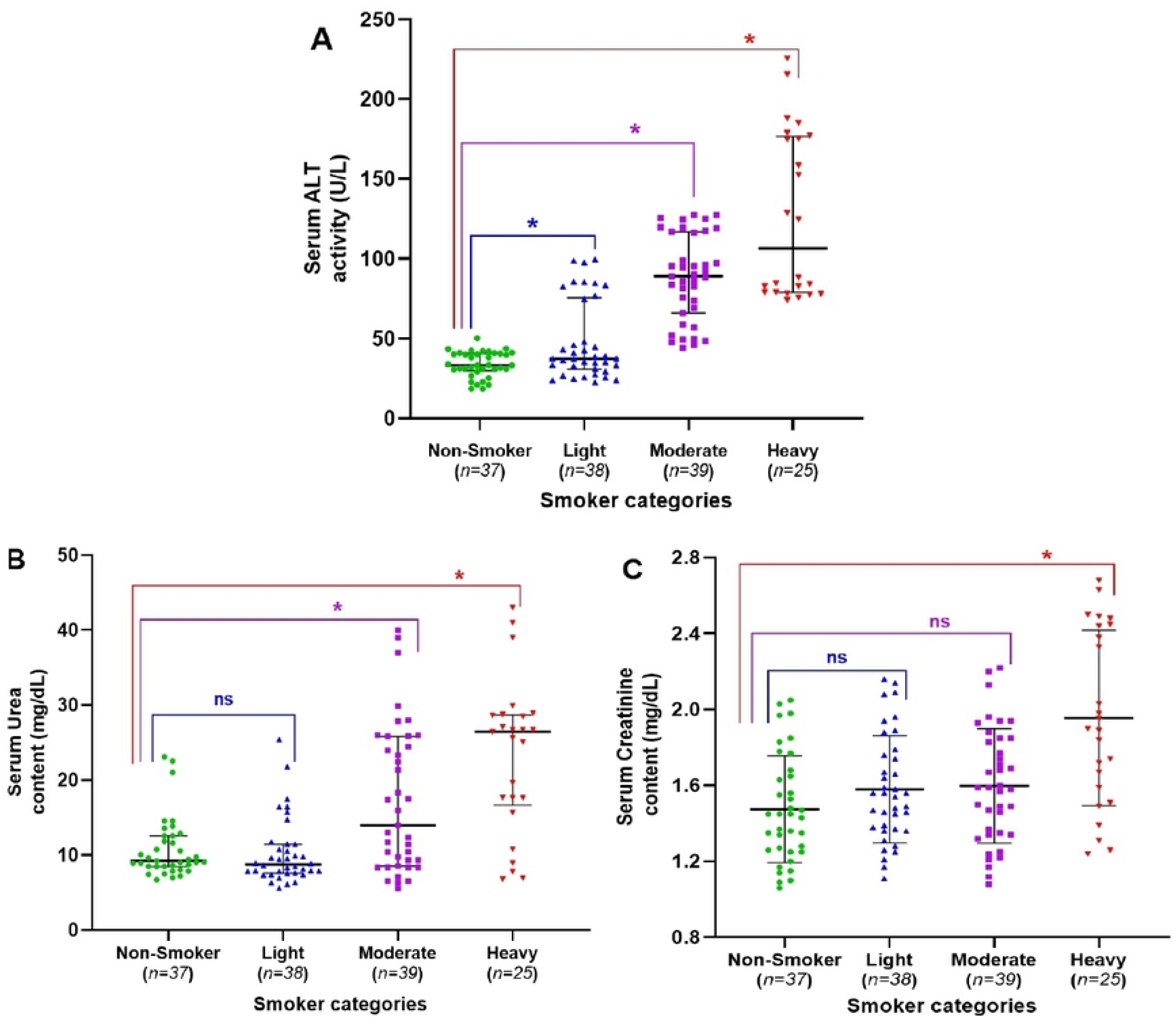
Variation of serum liver and kidney function markers per smoker categories ***(A):*** *Serum ALT activity**; (B):** Serum Urea content**; (C):** Serum Creatinine content. Values are expressed as median and interquartile ranges. **\***Median value significantly different when compared to Non-Smoker (P<0.05).* ***^ns^*** *Median value non-significantly different when compared to Non-Smoker (P˃0.05). ALT: Alanine aminotransferase*.

### Influence of potential confounding variables on variation of serum level of biochemical makers of liver and kidney functions among healthy active smokers

Sub-group analyses were performed among the enrolled active smokers to evaluate whether potential confounding variables including age, gender, duration of smoking, alcohol consumption, and electronic cigarette use could influence serum levels of ALT, Urea, and Creatinine. Regarding gender, serum ALT activity was significantly higher (P < 0.05) in males compared to females (Fig 3A), while no significant differences (P > 0.05) were observed between males and females for serum Urea (Fig 3B) or Creatinine levels (Fig 3C). Age groups did not significantly (P > 0.05) affect the levels of ALT (Fig 3D), Urea (Fig 3E), or Creatinine (Fig 3F).

**Fig 3:**
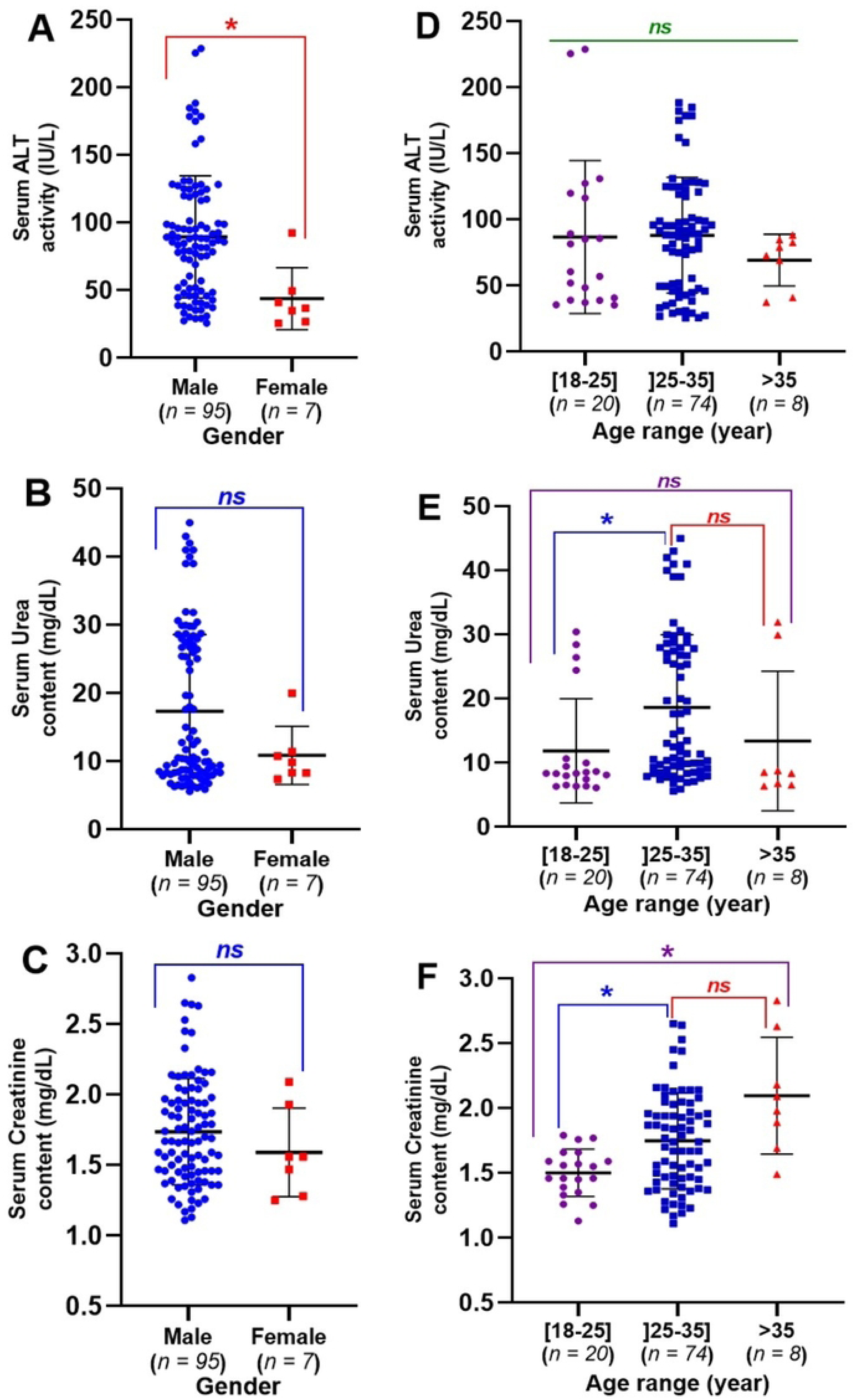
Sub-group analysis showing the influence of gender and age on the levels of serum markers of liver and kidneys damage in healthy smokers *(A) and (D), (B) and (E), (C) and (F): Influence of gender and age range on the variation of serum ALT activity, Urea and Creatinine contents, respectively. Values are expressed as median and interquartile ranges. *Median value significantly different between two compared groups (P<0.05). ^ns^ Median value non-significantly different between two compared groups (P˃0.05). ALT: Alanine aminotransferase*.

Similarly, the duration of smoking did not significantly (P > 0.05) influence the serum levels of ALT (Fig 4A), Urea (Fig 4B), or Creatinine (Fig 4C). In contrast, heavy and moderate smokers exhibited significantly higher (P < 0.05) levels of ALT (Fig 4D), Urea (Fig 4E), and Creatinine (Fig 4F) compared to light smokers. No significant (P > 0.05) differences in these parameters were observed between alcohol consumers and non-consumers, or between electronic cigarette users and non-users (Fig 5).

**Fig 4:**
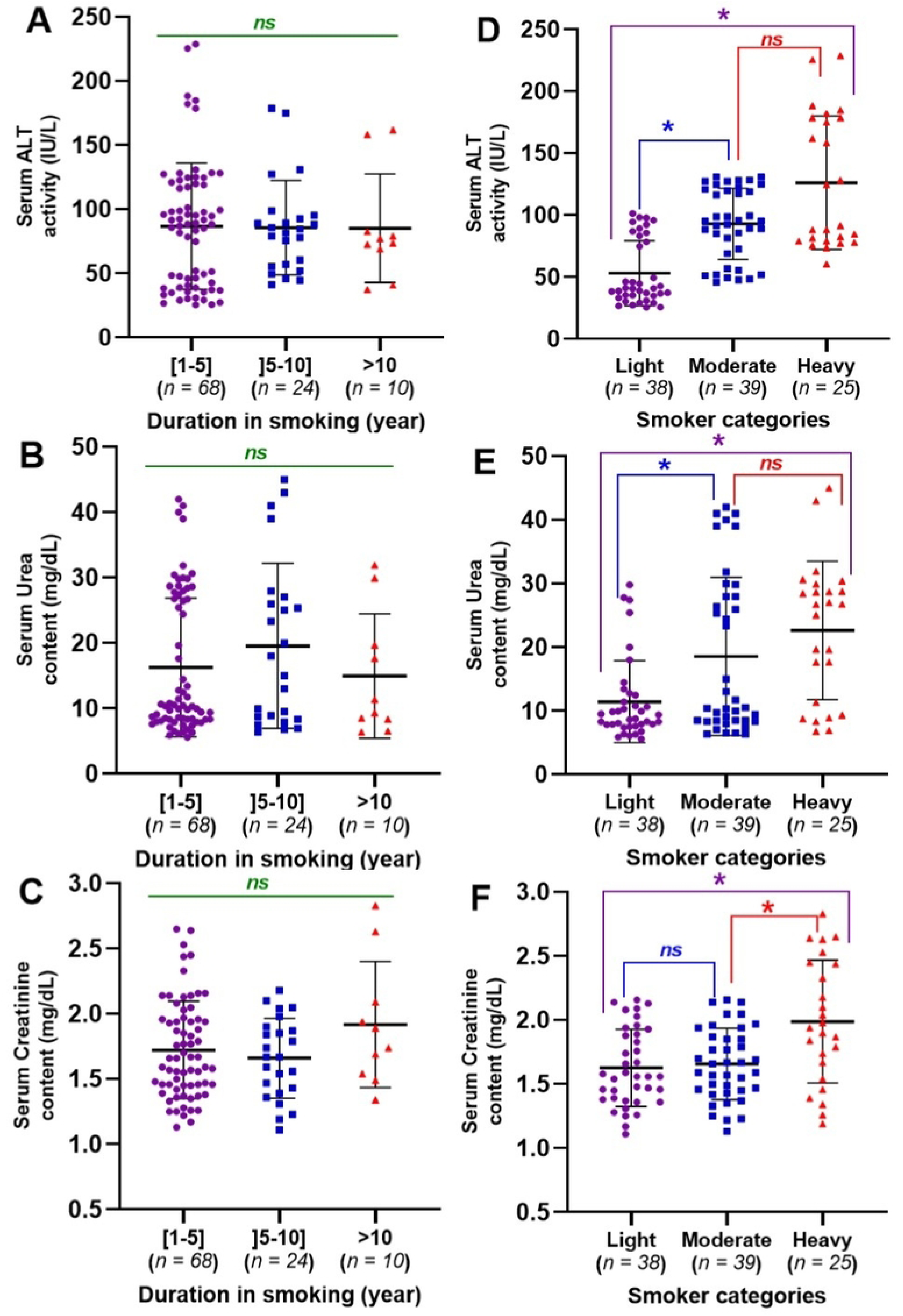
Sub-group analysis showing the influence of the duration in smoking and smoker categories on the levels of serum markers of liver and kidneys damage in healthy smokers *(A) and (D), (B) and (E), (C) and (F): Influence of the duration in smoking and smoker categories on the variation of serum ALT activity, Urea and Creatinine contents, respectively. Values are expressed as median and interquartile ranges. *Median value significantly different between two compared groups (P<0.05). ^ns^ Median value non-significantly different between two compared groups (P˃0.05). ALT: Alanine aminotransferase*.

**Fig 5:**
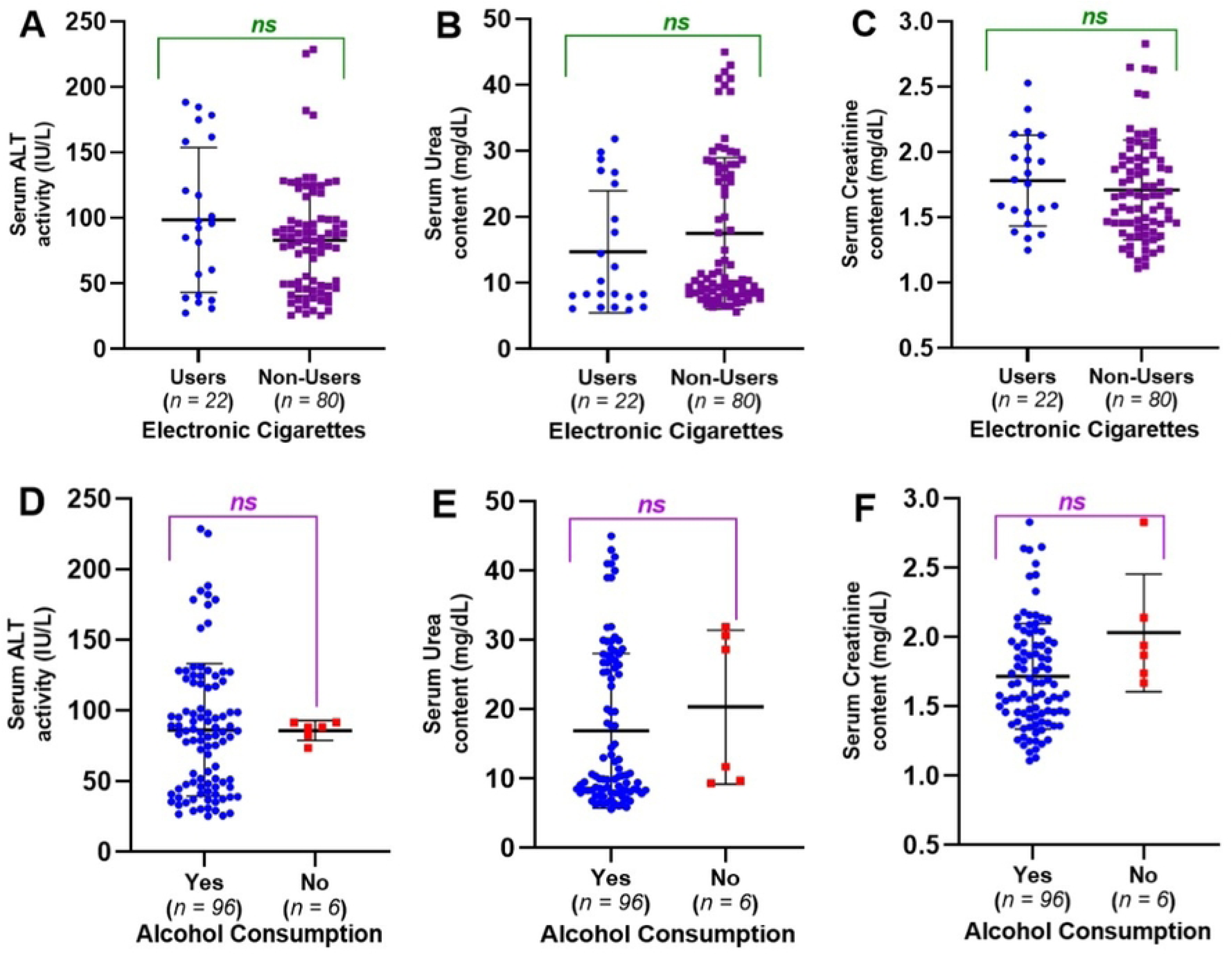
Sub-group analysis showing the influence of electronic cigarette and alcohol consumption on the levels of serum markers of liver and kidneys damage in healthy smokers *(A) and (D), (B) and (E), (C) and (F): Influence of the use of electronic cigarette and alcohol consumption on the variation of serum ALT activity, Urea and Creatinine contents, respectively. Values are expressed as median and interquartile ranges. *Median value significantly different between two compared groups (P<0.05). ^ns^ Median value non-significantly different between two compared groups (P˃0.05). ALT: Alanine aminotransferase*.

### Association between the smoker categories and the alteration of biochemical makers of liver and kidney function

The statistical associations between the smoker categories and the level of alterations of liver and kidney function markers are presented in Table 6. We observed that all smoker categories (light, moderate, and heavy) were at a significantly higher risk (RR range 4.86-13.28; *P*<0.05) of having abnormally high levels of serum ALT activity than the non-smokers. A similar result was obtained with the serum creatinine content (RR range 1.94-2.81; *P*<0.05). Light or moderate smokers were not at a significantly higher risk (RR range 1.02-2.57; *P*˃0.05) of having abnormal levels of serum content, positive albuminuria, and glucosuria than the non-smokers. However, heavy smokers were significantly at higher risk (RR range 6.90-13.32; *P*<0.05) of displaying abnormal elevated serum urea content, positive albuminuria, and glucosuria, when compared to the non-smokers.

**Table 6:** Statistical association between the variation of the liver and kidney function parameters and the smoker categories.

| Liver/kidneys function parameters |  | Smoker categories |  |  |  |
| --- | --- | --- | --- | --- | --- |
|  |  | Non-Smoker (37)<br>n (%) | Light (38)<br>n (%) | Moderate (39)<br>n (%) | Heavy (25)<br>n (%) |
| ALT<br>(IU/L) | High ($> 2 \times 40$) | 2 (5.4) | 10 (26.3) | 28 (71.7) | 16 (64.0) |
| | Normal ($< 2 \times 40$) | 35 (94.6) | 28 (73.7) | 11 (28.3) | 9 (36.0) |
|  | RR | 1.00 | **4.86** | **13.28** | **11.84** |
|  | [95% CI] | - | [1.3 – 19.0] | [3.9 – 48.5] | [3.4 – 43.8] |
|  | P-value | - | **0.0245\*** | **<0.0001\*** | **<0.0001\*** |
| Urea<br>(mg/dL) | High ($> 20$) | 3 (8.1) | 4 (10.5) | 16 (41.0) | 14 (56.0) |
| | Normal ($< 20$) | 34 (91.9) | 34 (89.5) | 23 (59.0) | 11 (44.0) |
|  | RR | 1.00 | 1.29 | **5.06** | **6.90** |
|  | [95% CI] | - | [0.3 – 4.9] | [1.7 – 15.4] | [2.4 – 20.9] |
| | P-value | - | $>0.9999$ | **0.0012\*** | **<0.0001\*** |
| Creatinine<br>(mg/dL) | High ($> 1.5$) | 10 (27.0) | 20 (52.6) | 27 (69.2) | 19 (76.0) |
| | Normal ($< 1.5$) | 27 (73.0) | 18 (47.4) | 12 (30.8) | 6 (24.0) |
|  | RR | 1.00 | **1.94** | **2.56** | **2.81** |
|  | [95% CI] | - | [1.08 – 3.6] | [1.5 – 4.6] | [1.6 – 5.1] |
|  | P-value | - | **0.0339\*** | **0.0003\*** | **0.0002\*** |
| Albuminuria | Positive | 2 (5.4) | 5 (13.1) | 20 (51.3) | 18 (72.0) |
|  | Negative | 35 (94.6) | 33 (86.9) | 19 (48.7) | 7 (28.0) |
|  | RR | 1.00 | 2.57 | **9.48** | **13.32** |
|  | [95% CI] | - | [0.5 – 10.2] | [2.7 – 35.2] | [3.9 – 48.9] |
|  | P-value | - | 0.4316 | **<0.0001\*** | **<0.0001\*** |
| Glucosuria | Positive | 3 (8.1) | 3 (7.9) | 4 (10.3) | 15 (60.0) |
|  | Negative | 34 (91.9) | 35 (92.1) | 35 (89.7) | 10 (40.0) |
|  | RR | 1.00 | 1.02 | 1.26 | **7.40** |
|  | [95% CI] | - | [0.2 – 4.01] | [0.3 – 4.8] | [2.6 – 22.2] |
| | P-value | - | $>0.9999$ | $>0.9999$ | **<0.0001\*** |

*The statistical association were performed using the Fisher Exact Test. Values of $P < 0.05$ were* *considered significantly different. RR: Relative Risk; CI: 95% Confidence Interval; Non-smokers* *(with $RR = 1.00$ ) were considered as reference during the analysis. RR and P-values highlighted in* *Bold indicate a significant association.*

### Association between potential confounders and the alteration of biochemical markers of kidney and liver damage among the enrolled healthy active smokers

Bivariate and multivariate logistic regression analyses were conducted to identify potential confounding variables such as age, gender, duration of smoking, alcohol consumption, and electronic cigarette use that could be associated with elevated levels of ALT, Urea, and Creatinine. The results indicated that gender (crude OR = 0.3; 95% CI: [0.15–1.14]; P = 0.066), age (cOR = 3.00; 95% CI: [0.48–18.60]; P = 0.238), alcohol consumption (cOR = 1.84; 95% CI: [0.32–10.53]; P = 0.493), and electronic cigarette use (cOR = 0.57; 95% CI: [0.21–1.51]; P = 0.260) were not significantly associated with high serum ALT levels (Table 7). Similar patterns were observed for abnormal serum Urea levels (Table 8) and Creatinine levels (Table 9). In contrast, both bivariate and multivariate analyses confirmed that smoking duration and the number of cigarettes smoked per day (smoker categories) were significantly associated with elevated serum ALT levels in healthy active smokers (Table 7). Individuals with less than five years of smoking history had a significantly lower risk (adjusted OR = 16.07; 95% CI: [2.45–105.39]; P = 0.004) compared to those with more than five years of smoking. Similarly, light smokers showed a significantly reduced risk of elevated serum ALT (aOR = 0.11; 95% CI: [0.038–0.32]; P < 0.0001) compared to moderate or heavy smokers (Table 7). These findings were also reflected in serum Urea levels (Table 8). However, no significant associations were observed between smoking history or smoker categories and serum Creatinine levels (Table 9).

**Table 7:** Bivariate and multivariate logistic regression analysis for the identification of potential confounders associated with the high serum level of ALT among enrolled smokers.

| Confounding variables | Categories | ALT level |  |  | Bivariate logistic regression |  |  | Multivariate logistic regression |  |  |
| --- | --- | --- | --- | --- | --- | --- | --- | --- | --- | --- |
|  |  | High n, (%) | Normal n, (%) | Total | cOR | [95% CI] | P values | aOR | [95% CI] | P values |
| Gender | Female | 1 (1) | 6 (5.9) | 7 (6.9) | 0.13 | [0.15 – 1.14] | 0.066 |  |  |  |
|  | Male | 53 (52.0) | 42 (41.2) | 95 (93.1) | 1 | / | / |  | / |  |
|  | Total | 54 (52.9) | 48 (47.1) | 102 (100.0) | / | / | / |  |  |  |
| Age range (year) | [18-25] | 10 (9.8) | 10 (9.8) | 20 (19.6) | 3.00 | [0.48 – 18.60] | 0.238 |  |  |  |
|  | ]25-35] | 42 (41.2) | 32 (31.4) | 74 (72.5) | 3.94 | [0.74 – 20.81] | 0.107 |  | / |  |
|  | >35 | 2 (2.0) | 6 (5.9) | 8 (7.8) | 1 | / | / |  |  |  |
|  | Total | 54 (52.9) | 48 (47.1) | 102 (100.0) | / | / | / |  |  |  |
| Duration in smoking (Year range) | [1-5] | 40 (39.2) | 28 (27.5) | 68 (66.7) | 5.71 | [1.12 – 28.96] | 0.035* | 16.07 | [2.45 – 105.39] | 0.004* |
|  | ]5-10] | 12 (11.8) | 12 (11.8) | 24 (23.5) | 4.00 | [0.69 – 22.87] | 0.119 | 5.63 | [0.87 – 36.49] | 0.070 |
|  | >10 | 2 (2.0) | 8 (7.8) | 10 (9.8) | 1 | / | / | 1 | / | / |
|  | Total | 54 (52.9) | 48 (47.1) | 102 (100.0) | / | / | / | / | / | / |
| Smoker category | Light | 10 (9.8) | 28 (27.5) | 38 (37.3) | 0.14 | [0.05 – 0.38] | <0.0001* | 0.11 | [0.038 – 0.32] | <0.0001* |
|  | Moderate | 28 (27.5) | 11 (10.8) | 39 (38.2) | 1 | / | / | 1 | / | / |
|  | Heavy | 16 (15.7) | 9 (8.8) | 25 (24.5) | 0.39 | [0.23 – 2.04] | 0.512 | 1.41 | [0.39 – 5.07] | 0.598 |
|  | Total | 54 (52.9) | 48 (47.1) | 102 (100.0) | / | / | / |  |  |  |
| Alcohol consumption | No | 4 (3.9) | 2 (2.0) | 6 (5.9) | 1.84 | [0.32 – 10.53] | 0.493 |  |  |  |
|  | Yes | 50 (49.0) | 46 (45.1) | 96 (94.1) | 1 | / | / |  | / |  |
|  | Total | 54 (52.9) | 48 (47.1) | 102 (100.0) | / | / | / |  |  |  |
| Use of E-cigarette | No | 40 (39.2) | 40 (39.2) | 80 (78.4) | 0.57 | [0.21 – 1.51] | 0.260 |  |  |  |
|  | Yes | 14 (13.7) | 8 (7.8) | 22 (21.6) | 1 | / | / |  | / |  |
|  | Total | 54 (52.9) | 48 (47.1) | 102 (100.0) | / | / | / |  |  |  |

E-cigarette: Electronic cigarette; cOR: Crude Odd Ratio; aOR: Adjusted Odd Ratio; CI: Confidence Interval; The bold cOR or aOR and P-value are indicators of a significant association. \* Indicates $P < 0.05$ . Categorical variables with cOR or aOR = 1.00 were automatically considered by the software as reference during the analysis.

**Table 8:**
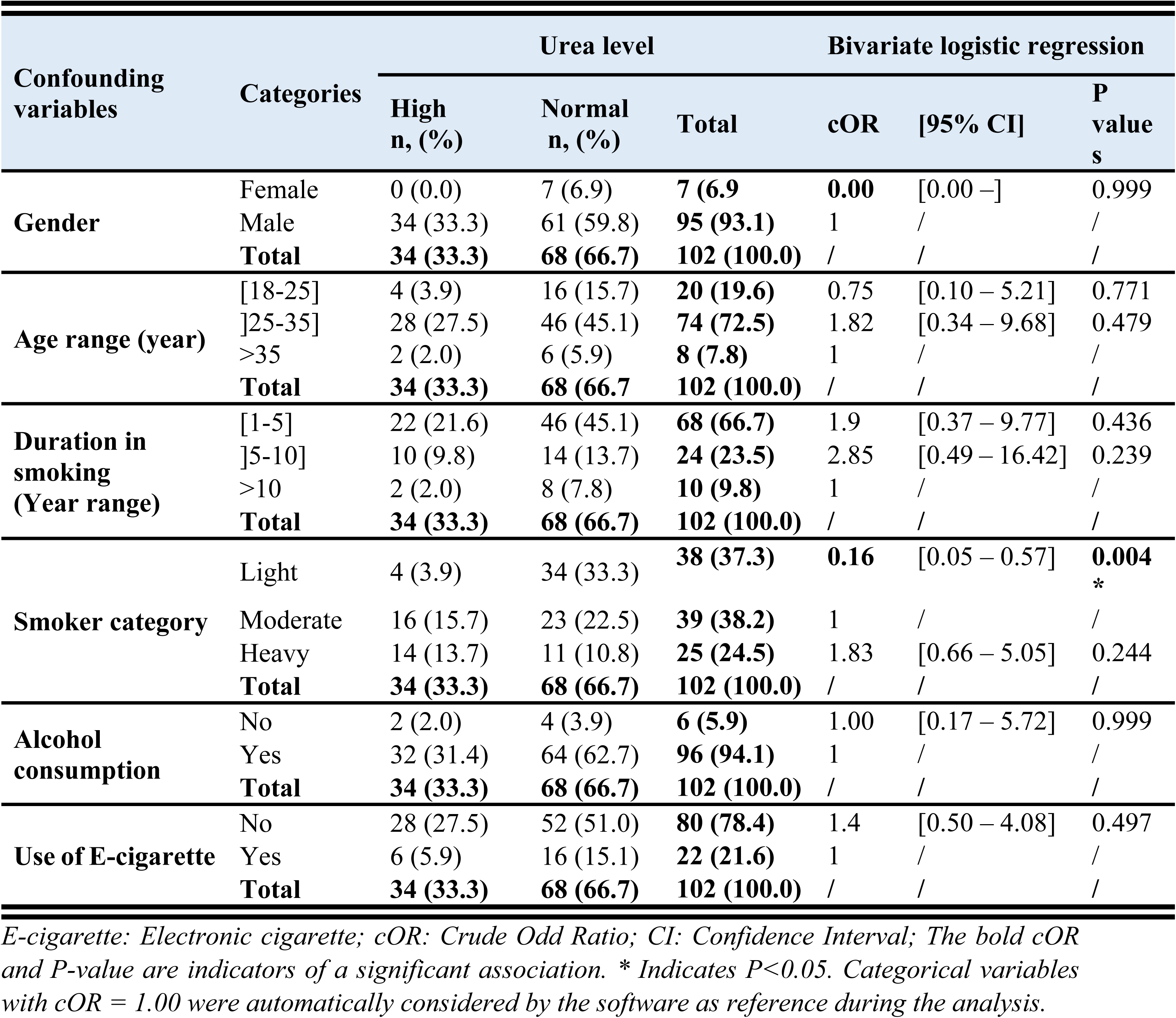
Bivariate logistic regression analysis for the identification of potential confounders associated with the high serum level of urea among enrolled smokers.

**Table 9:** Bivariate logistic regression analysis for the identification of potential confounders associated with the high serum level of creatinine among enrolled smokers.

| Confounding variables | Categories | Creatinine level |  |  | Bivariate logistic regression |  |  |
| --- | --- | --- | --- | --- | --- | --- | --- |
|  |  | High n, (%) | Normal n, (%) | Total | cOR | [95% CI] | P values |
| Gender | Female | 3 (2.9) | 4 (3.9) | **7 (6.9)** | 0.38 | [0.80 – 1.80] | 0.224 |
|  | Male | 63 (61.8) | 32 (31.4) | **95 (93.1)** | 1 | / | / |
|  | **Total** | **66 (64.7)** | **36 (35.3)** | **102 (100.0)** | / | / | / |
| Age range (year) | [18-25] | 10 (9.8) | 10 (9.8) | **20 (19.6)** | 0.00 | [0.00 –] | 0.999 |
|  | ]25-35] | 48 (47.1) | 26 (25.5) | **74 (72.5)** | 0.00 | [0.00 –] | 0.999 |
|  | >35 | 8 (7.8) | 0 (0.0) | **8 (7.8)** | 1 | / | / |
|  | **Total** | **66 (64.7)** | **36 (35.3)** | **102 (100.0)** | / | / | / |
| Duration in smoking (Year range) | [1-5] | 42 (41.2) | 26 (25.5) | **68 (66.7)** | 0.40 | [0.80 – 2.05] | 0.274 |
|  | ]5-10] | 16 (15.7) | 8 (7.8) | **24 (23.5)** | 0.50 | [0.85 – 2.92] | 0.442 |
|  | >10 | 8 (7.8) | 2 (2.0) | **10 (9.8)** | 1 | / | / |
|  | **Total** | **66 (64.7)** | **36 (35.3)** | **102 (100.0)** | / | / | / |
| Smoker category | Light | 20 (19.6) | 18 (17.6) | **38 (37.3)** | 0.49 | [0.19 – 1.25] | 0.138 |
|  | Moderate | 27 (26.5) | 12 (11.8) | **39 (38.2)** | 1 | / | / |
|  | Heavy | 19 (18.6) | 6 (5.9) | **25 (24.5)** | 1.40 | [0.44 – 4.41] | 0.558 |
|  | **Total** | **66 (64.7)** | **36 (35.3)** | **102 (100.0)** | / | / | / |
| Alcohol consumption | No | 6 (5.9) | 0 (0.0) | **6 (5.9)** | 0.00 | [0.00 –] | 0.999 |
|  | Yes | 60 (58.8) | 36 (35.3) | **96 (94.1)** | 1 | / | / |
|  | **Total** | **66 (64.7)** | **36 (35.3)** | **102 (100.0)** | / | / | / |
| Use of E-cigarette | No | 52 (51.0) | 28 (27.5) | **80 (78.4)** | 1.06 | [0.39 – 2.38] | 0.906 |
|  | Yes | 14 (13.7) | 8 (7.8) | **22 (21.6)** | 1 | / | / |
|  | **Total** | **66 (64.7)** | **36 (35.3)** | **102 (100.0)** | / | / | / |

*E-cigarette: Electronic cigarette; cOR: Crude Odd Ratio; CI: Confidence Interval; The bold cOR* *and P-value are indicators of a significant association. \* Indicates P<0.05. Categorical variables* *with cOR = 1.00 were automatically considered by the software as reference during the analysis.*

## Discussion

Smoking cigarettes has disastrous impacts on human health, resulting in over 8 million deaths directly and being a contributing factor in over 1.2 million deaths globally, every year [9]. Cigarettes have numerous harmful substances like cadmium which can harm the lungs and also damage the liver and kidneys, two crucial organs responsible for detoxifying the body [13,29]. There is a concerning rise in smoking among young adults in developing nations, and Cameroon is no different. Regrettably, in these nations, there is a lack of information on the risk exposure to toxic substances like cadmium from smoking, as well as the impact of smoking on the liver and kidney functions of these young individuals. Accordingly, this research was undertaken to evaluate how smoking cigarettes affects the biochemical indicators of liver and kidney damage in healthy active smokers in Buea, Cameroon. Additionally, an estimation of cadmium risk exposure through smoking based on the most popular brands of cigarettes consumed in Buea was also performed.

Our findings show that most of the individuals who took part in the study are young adult males (93.1%), ranging in age from 25 to 35 years old (72.5%). On average, they smoked 10 cigarettes per day and were categorized as either moderate (38.2%) or heavy (24.5%) smokers, with a monthly income of less than 100,000 CFA francs (less than 200 US Dollars). With the addictive nature of smoking taken into account, it is anticipated that young adults who continue to smoke without cessation will experience limited personal socio-economic growth and negative health consequences in the future. All brands of cigarettes identified from the survey were of foreign origin and the analysis confirmed the bioaccumulation of cadmium in tobacco leaves, which represent the primary raw material in the cigarette industry [8]. Indeed, the concentration of cadmium in the identified cigarette brands consumed in Buea ranged from 0.72 ± 0.07 to 1.08 ± 0.09 µg Cd/g of cigarette. The cadmium levels in these products are comparable to those reported in studies on cadmium content in cigarettes from different countries worldwide [6,8,24]. These results also indicate that smoking cigarettes, regardless of the brand, consistently exposes the smoker to cadmium and its harmful effects on health.

Considering the long half-life of cadmium in the human body (between 16-30 years) [30], it was important to estimate the Daily Exposure (DE) and Weekly Inhalational Exposure (WIE) to cadmium and its percentage contribution to the Tolerable Weekly Intake (%TWI) of cadmium from smoking. The findings showed that Cadmium DE ranged from 0.18 to 1.44 µg Cd/day, with smoker patterns like cigarette brand and the daily number of cigarettes consumed affecting the levels. Compared to the permitted No Significant Risk Level (NSRL) – Inhalation of cadmium established at 0.05 µg Cd/day by the California Environmental Protection Agency [25], these findings indicated that heavy smokers in Buea are up to 28-fold more exposed to cadmium via inhalation than non-smokers. Likewise, our data indicated that cigarette smoking represented 4-8% of the TWI of cadmium. Even though these relative contributions appear to be small, when considering other sources of cadmium inhalation such as environmental and occupational exposure, the actual contribution of the WIE to the TWI of cadmium could be considerably increased. These findings indicate worry for both heavy smokers and those who smoke less than 5 cigarettes a day when considering other sources of cadmium exposures such as inhalation of industrial emissions [31]. Thus, there is a high likelihood that the total TWI of cadmium in healthy active smokers in Buea may exceed the permitted TWI of 2.5 µg Cd/kg b.w. per week, set by the European Food Safety Authority [26], if all sources and routes of contamination are considered.

Cigarette smoking is widely recognized as a significant risk factor for numerous serious illnesses including respiratory, cardiovascular, and nervous system diseases (Jain and James, 2013). Furthermore, multiple studies have shown that smoking can result in chronic liver and kidney diseases over time, as well as change the normal levels of biochemical markers for kidney and liver functions, indicating the early stages of damage to these organs [18,32,33]. The results from our research indicated that levels of serum ALT activity were significantly higher (*P*<0.05) in all groups of smokers when compared to non-smokers and that heavy and moderated smokers also displayed a significant (P<0.05) higher level of serum ALT activity, as compared to light smokers. The relative risk for elevated serum ALT levels in light, moderate, and heavy smokers compared to non-smokers were 4.86, 13.28, and 11.84, respectively. Given that ALT is a cytosolic enzyme that is plentiful in the cytosol of hepatocytes, its abnormally increased levels in the bloodstream may suggest hepatocyte necrosis [12,34–36]. These results indicate that smoking may directly harm the liver, causing damage to hepatocytes and leading to higher levels of serum ALT activity in smokers, and that the severity of the damage increases with the number of cigarettes smoked per day.

Concerning the kidney function tests, our findings showed a significant (P<0.05) increase in serum urea and creatinine contents among healthy active smokers compared to non-smokers, indicating potential early signs of kidney function impairment. Sub-group analysis also revealed that heavy and moderate smokers displayed a significant (P<0.05) high level of Urea and Creatinine, when compared to light smokers. This finding also suggests that the severity of the impact of cigarette smoking on the kidneys increases with the quantity of cigarette smoked per day. Likewise, we observed that albuminuria and glucosuria tests showed positive results, particularly among moderate and heavy smokers, who had a respective RR of 7.40 and 13.32 of having positive test results compared to non-smokers. Given that glucose is expected to be completely reabsorbed in the proximal tubule of the kidney [37], these findings could indicate early signs of reduced reabsorption capacity in the proximal tubule of moderate and heavy smokers’ kidneys. Similarly, the elevated prevalence of urinary albuminuria, which is predominantly found in moderate (51.3%) and heavy (72.0%) smokers as opposed to non-smokers (5.4%), could indicate glomerular damage as a result of smoking cigarettes. These findings indicate that smokers face a greater likelihood of developing chronic kidney diseases when compared to non-smokers. These results align with prior studies analyzing urine samples from 140 active smokers without known kidney disease symptoms, showing a high albuminuria prevalence (75.7%) compared to controls (2.1%) [33], confirming the impact of smoking on the kidneys in healthy smokers.

This study has several potential limitations. First, regarding gender representativity, participants were predominantly male (93.1%), which reflects the overall smoking population, as smoking is more prevalent among men. Also, recruitment was based on voluntary consent, and fewer potential female participants were reluctant to participate, resulting in a limited number of females. Consequently, this imbalance may affect the validity of the findings across both sexes. Future research should aim to recruit more female participants to enhance the representativeness and validity of the results for both genders. Second, the estimation of cadmium exposure was based solely on the cadmium content in cigarettes and smoking behaviors. It did not include biological monitoring, such as urinary cadmium levels, which would better account for individual variability in absorption, metabolism, and accumulation of cadmium. Our results indicated that smokers could be up to 28 times more exposed to cadmium via inhalation than non-smokers, and that smoking contributes approximately 8% of the Tolerable Weekly Intake (%TWI) of cadmium. It is plausible that urinary cadmium concentrations are higher in smokers compared to non-smokers, as previous studies have demonstrated significant increases in urinary cadmium among smokers [17,38]. Further research is necessary to validate these findings within the Cameroon context and to consider other sources and routes of cadmium exposure. Additionally, some potential confounders that could influence liver and kidney biomarkers were evaluated. Our results showed no significant associations between these biomarkers and variables such as alcohol consumption, gender, or age. However, this may be due to the underrepresentation of certain sub-groups—for example, females and non-alcohol consumers comprised only 6.9% and 5.9% of participants, respectively. Therefore, these findings cannot be generalized to the broader community. Future studies with balanced representation of all sub-groups are needed to confirm the current findings.

## Conclusion

The current investigation involved determining the level of cadmium inhalational exposure through smoking, along with the association of cigarette smoking with the abnormal level of liver and kidney function biomarkers in healthy active smokers of the city of Buea, Cameroon. Our results demonstrated that active smokers are highly exposed to cadmium inhalation, up to 28-fold beyond the permitted No Significant Risk Level—inhalation of cadmium established at 0.05 µg Cd/day; and are also at higher risk of displaying abnormally increased levels of biochemical markers of liver and kidney functions, suggesting the occurrence of early signs of organs damage. Overall, these findings highlight the implications of cigarette smoking and high levels of cadmium exposure on the health risks associated with renal and hepatic diseases in apparently healthy active smokers. Based on the conclusion of this investigation, we advise smokers to give up smoking due to its harmful impact on the body. When it is difficult to quit, we suggest reducing the number of cigarettes smoked daily. It is also advocate switching to e-cigarettes when nicotine addiction makes quitting difficult. The harm reduction model proposes that using e-cigarettes can significantly reduce exposure to carcinogens and cadmium, as they deliver nicotine without burning tobacco leaves. We also recommend regularly monitoring the biochemical markers of liver and kidney functions for all active smokers given that it could be essential to prevent additional harm to these organs. Finally, we suggest the authorities and law enforcement officers to increase advertising and awareness efforts targeted at discouraging the public from smoking.

## Data Availability

All relevant data are within the manuscript and its Supporting Information files.

## Acknowledgment

The authors are grateful to all the participants who took part in this investigation.

## Supporting information

S1_file: Questionnaire

S2_file: Research Data

## References

1. Smereczański NM, Brzóska MM. Current Levels of Environmental Exposure to Cadmium in Industrialized Countries as a Risk Factor for Kidney Damage in the General Population: A Comprehensive Review of Available Data. Int J Mol Sci. 2023;24: 8413. doi:10.3390/ijms24098413

2. Mérida-Ortega Á, López-Carrillo L, Rangel-Moreno K, Ramirez N, Rothenberg SJ. Tobacco Smoke Exposure and Urinary Cadmium in Women from Northern Mexico. Int J Environ Res Public Health. 2021;18: 12581. doi:10.3390/ijerph182312581

3. Ganguly K, Levänen B, Palmberg L, Åkesson A, Lindén A. Cadmium in tobacco smokers: a neglected link to lung disease? Eur Respir Rev Off J Eur Respir Soc. 2018;27: 170122. doi:10.1183/16000617.0122-2017

4. Benson NU, Anake WU, Adedapo AE, Fred-Ahmadu OH, Ayejuyo OO. Toxic metals in cigarettes and human health risk assessment associated with inhalation exposure. Environ Monit Assess. 2017;189: 619. doi:10.1007/s10661-017-6348-x

5. Sandal S, Verghese PS, Taneja A, Massey DD, Habil M. Cigarettes as a source of heavy metal toxicity: evaluating human health risks. Discov Public Health. 2025;22: 311. doi:10.1186/s12982-025-00650-2

6. Caruso R, O’Connor R, Stephens W, Cummings K, Fong G. Toxic Metal Concentrations in Cigarettes Obtained from U.S. Smokers in 2009: Results from the International Tobacco Control (ITC) United States Survey Cohort. Int J Environ Res Public Health. 2013;11: 202–217. doi:10.3390/ijerph110100202

7. Lugon-Moulin N, Martin F, Krauss MR, Ramey PB, Rossi L. Cadmium concentration in tobacco (*Nicotiana tabacum* L.) from different countries and its relationship with other elements. Chemosphere. 2006;63: 1074–1086. doi:10.1016/j.chemosphere.2005.09.005

8. Genchi G, Sinicropi MS, Lauria G, Carocci A, Catalano A. The Effects of Cadmium Toxicity. Int J Environ Res Public Health. 2020;17: 3782. doi:10.3390/ijerph17113782

9. WHO. Tobacco: who.int/news-room/fact-sheets/detail/tobacco. 2023 [cited 4 Oct 2024]. Available: https://www.who.int/news-room/fact-sheets/detail/tobacco

10. Njoumemi Z, Fadimatou A, Bwemba N. Prevalence and Socioeconomic Determinants of Tobacco Consumption Patterns across Urban and Rural Settings in Cameroon. J US-China Med Sci. 2020;17. doi:10.17265/1548-6648/2020.04.007

11. Frey R, Becker C, Unger S, Schmidt A, Wensing G, Mück W. Assessment of the effects of renal impairment and smoking on the pharmacokinetics of a single oral dose of the soluble guanylate cyclase stimulator riociguat (BAY 63-2521). Pulm Circ. 2016;6: S15–S26. doi:10.1086/685017

12. Kouam AF, Masso M, Kouoh FE, Fifen R, Njingou I, Tchana AN, et al. Hydro-ethanolic extract of Khaya grandifoliola attenuates heavy metals-induced hepato-renal injury in rats by reducing oxidative stress and metals-bioaccumulation. Heliyon. 2022;8: e11685. doi:10.1016/j.heliyon.2022.e11685

13. El-Zayadi A-R. Heavy smoking and liver. World J Gastroenterol. 2006;12: 6098. doi:10.3748/wjg.v12.i38.6098

14. Muriel P. Role of free radicals in liver diseases. Hepatol Int. 2009;3: 526–536. doi:10.1007/s12072-009-9158-6

15. Marti-Aguado D, Clemente-Sanchez A, Bataller R. Cigarette smoking and liver diseases. J Hepatol. 2022;77: 191–205. doi:10.1016/j.jhep.2022.01.016

16. Abdel-Rahman O, Helbling D, Schöb O, Eltobgy M, Mohamed H, Schmidt J, et al. Cigarette smoking as a risk factor for the development of and mortality from hepatocellular carcinoma: An updated systematic review of 81 epidemiological studies. J Evid-Based Med. 2017;10: 245–254. doi:10.1111/jebm.12270

17. Mortensen ME, Wong L-Y, Osterloh JD. Smoking status and urine cadmium above levels associated with subclinical renal effects in U.S. adults without chronic kidney disease. Int J Hyg Environ Health. 2011;214: 305–310. doi:10.1016/j.ijheh.2011.03.004

18. Yardimci B, Ecder T. Smoking and Chronic Kidney Disease. Turk J Nephrol. 2019;28: 75–80. doi:10.5152/turkjnephrol.2019.3440

19. Yan L-J, Allen DC. Cadmium-Induced Kidney Injury: Oxidative Damage as a Unifying Mechanism. Biomolecules. 2021;11: 1575. doi:10.3390/biom11111575

20. Hong D, Min J-Y, Min K-B. Association Between Cadmium Exposure and Liver Function in Adults in the United States: A Cross-sectional Study. J Prev Med Pub Health. 2021;54: 471. doi:10.3961/jpmph.21.435

21. Mapa-Tassou C, Bonono CR, Assah F, Wisdom J, Juma PA, Katte J-C, et al. Two decades of tobacco use prevention and control policies in Cameroon: results from the analysis of non-communicable disease prevention policies in Africa. BMC Public Health. 2018;18: 958. doi:10.1186/s12889-018-5828-4

22. Cochran WG. Sampling techniques, Third Edition. John Wiley & Sons. 1977 [cited 13 Aug 2024]. Available: https://www.academia.edu/29684662/Cochran_1977_Sampling_Techniques_Third_Edition

23. Njoumemi Z, Fadimatou A, Bwemba N. Prevalence and Socioeconomic Determinants of Tobacco Consumption Patterns across Urban and Rural Settings in Cameroon. J US-China Med Sci. 2020;17. doi:10.17265/1548-6648/2020.04.007

24. Vlachou C, Vejdovszky K, Wolf J, Steinwider J, Fuchs K, Hofstädter D. Toxicological approaches for the quantitative inhalation risk assessment of toxic metals from tobacco smoke: application on the deterministic and probabilistic inhalation risk assessment of cadmium for Austrian smokers. Inhal Toxicol. 2021;33: 128–142. doi:10.1080/08958378.2021.1912859

25. CalEPA-OEHHA. Cadmium: No Significant Risk Level (NSRL) - Inhalation. (California Environmental Protection AgencyOffice of Environmental Health Hazard Assessment). In: OEHHA [Internet]. 2015 [cited 4 Oct 2024]. Available: https://oehha.ca.gov/chemicals/cadmium

26. EFSA EFS. Cadmium dietary exposure in the European population: European Food Safety Authority. EFSA J. 2012;10: 2551. doi:10.2903/j.efsa.2012.2551

27. Essam MYN, Kouam AF, Fepa AGK, Seukep AJ, Zeuko’o EM, Somene FSD, et al. Serological evidence and factors associated to liver damage in malaria-typhoid infected patients consulting in two health facilities, Yaoundé-Cameroon. PLOS ONE. 2025;20: e0319547. doi:10.1371/journal.pone.0319547

28. NIH. Roussel Uclaf Causality Assessment Method (RUCAM) in Drug Induced Liver Injury. LiverTox: Clinical and Research Information on Drug-Induced Liver Injury. Bethesda (MD): National Institute of Diabetes and Digestive and Kidney Diseases; 2012. Available: http://www.ncbi.nlm.nih.gov/books/NBK548272/

29. Lang SM, Schiffl H. Smoking status, cadmium, and chronic kidney disease. Ren Replace Ther. 2024;10: 17. doi:10.1186/s41100-024-00533-3

30. Charkiewicz AE, Omeljaniuk WJ, Nowak K, Garley M, Nikliński J. Cadmium Toxicity and Health Effects—A Brief Summary. Molecules. 2023;28: 6620. doi:10.3390/molecules28186620

31. Ruczaj A, Brzóska MM. Environmental exposure of the general population to cadmium as a risk factor of the damage to the nervous system: A critical review of current data. J Appl Toxicol JAT. 2023;43: 66–88. doi:10.1002/jat.4322

32. Bandiera S, Pulcinelli RR, Huf F, Almeida FB, Halmenschlager G, Bitencourt PER, et al. Hepatic and renal damage by alcohol and cigarette smoking in rats. Toxicol Res. 2021;37: 209–219. doi:10.1007/s43188-020-00057-y

33. Eid HA, Moazen EM, Elhussini M, Shoman H, Hassan A, Elsheikh A, et al. The Influence of Smoking on Renal Functions Among Apparently Healthy Smokers. J Multidiscip Healthc. 2022;Volume 15: 2969–2978. doi:10.2147/JMDH.S392848

34. Kouam AF, Mofor SM, Essam MYN, Fepa AGK, Zeuko’o EM, Seukep AJ, et al. Abnormal serum levels of liver enzyme markers and related risk factors in type 2 diabetes mellitus patients attending the Buea Regional Hospital, Cameroon. Tian S, editor. PLOS One. 2025;20: e0328974. doi:10.1371/journal.pone.0328974

35. Kouam AF, Ngoumé NAN, Fepa AGK, Wainfen Z, Ngounou E, Galani BRT, et al. Liver injury in malaria infected patients in Douala-Cameroon and its association with poor medical practice. Egypt Liver J. 2023;13: 67. doi:10.1186/s43066-023-00300-9

36. Qin S, Wang J, Yuan H, He J, Luan S, Deng Y. Liver function indicators and risk of hepatocellular carcinoma: a bidirectional mendelian randomization study. Front Genet. 2024;14. doi:10.3389/fgene.2023.1260352

37. Blanchard A, Poussou R, Houillier P. Exploration des fonctions tubulaires rénales. Néphrologie Thérapeutique. 2009;5: 68–83. doi:10.1016/j.nephro.2008.03.004

38. Ikeda M, Moriguchi J, Ezaki T, Fukui Y, Ukai H, Okamoto S, et al. Smoking-induced increase in urinary cadmium levels among Japanese women. Int Arch Occup Environ Health. 2005;78: 533–540. doi:10.1007/s00420-005-0612-z

